# Measuring up: A Comparison of Tapestation 4200 and Bioanalyzer 2100 as Measurement Tools for RNA Quality in Postmortem Human Brain Samples

**DOI:** 10.1101/2023.07.03.23291969

**Authors:** Jessica E Walker, Javon C Oliver, Analisa M Stewart, Suet Theng Beh, Richard A Arce, Michael J Glass, Daisy E Vargas, Sanaria H Qiji, Anthony J Intorcia, Claryssa I Borja, Madison P Cline, Spencer J Hemmingsen, Addison N Krupp, Rylee D McHattie, Monica R Mariner, Ileana Lorenzini, Sidra Aslam, Cecilia Tremblay, Thomas G Beach, Geidy E Serrano

## Abstract

Determining RNA integrity is a critical quality assessment tool for gene expression studies where the experiment’s success is highly dependent on sample quality. Since its introduction in 1999, the gold standard in the scientific community has been the Agilent 2100 Bioanalyzer’s RNA Integrity Number (RIN) which uses a 1-10 value system with 1 being the most degraded to 10 being the most intact. In 2015, Agilent launched the 4200 Tapestation’s RIN equivalent and reported a strong correlation of r^2^ of 0.936 and median error < ± 0.4 RIN units. To evaluate this claim, we compared the Agilent 4200 Tapestation’s RIN equivalent (RINe) and DV200 to the Agilent 2100 Bioanalyzer’s RIN for 183 parallel RNA samples. In our study, using RNA from a total of 183 human postmortem brain samples, we found that the RIN and RINe values only weakly correlate with an r^2^ of 0.393 and an average difference of 3.2 RIN units. DV200 also only weakly correlated with RIN (r^2^ of 0.182) and RINe (r^2^ of 0.347). Finally, when applying a cut-off value of 6.5 for both metrics, we found that 95.6% of samples passed with RIN, while only 23.5% passed with RINe. Our results suggest that even though RIN (Bioanalyzer) and RINe (Tapestation) use the same 1-10 value system, they should not be used interchangeably, and cut-off values should be calculated independently.

## Introduction

The RIN (RNA Integrity Number) measured on the Agilent 2100 Bioanalyzer is widely accepted in the scientific community as the gold standard for objectively assessing a sample’s RNA quality for downstream applications. The Bioanalyzer is an automated microfluidics-based platform that separates RNA by molecular weight and detects RNA using a fluorescent intercalated dye and fluorescent detecting laser. The results are displayed in a gel-like image for visualizing fragment size and distribution, accompanied by an electropherogram (peaks) for RIN visualization based on a RIN value of 1 (totally degraded) to 10 (fully intact). A proprietary algorithm is assigned to the sample, reportedly making RIN easily reproducible between users (Schroeder et al., 2006).

Previously, RNA quality was evaluated by performing gel electrophoresis followed by analyzing the 28S:18S ribosomal bands where RNA with a 28S:18S ratio of 2 or greater is considered to be of high quality. This method is still used but is sample-demanding (0.5-2ug total RNA) and dependent on subjective human interpretation (Imbeaud et al., 2005). Since the Bioanalyzer was first introduced in 1999, scientists have been using RIN values to identify acceptable samples for specific downstream applications For example, RIN > 6.5 is considered acceptable for most gene expression assays (Kap, Oomen, Arshad, de Jong, & Riegman, 2014).

In recent years, new instruments for measuring RNA integrity have been developed. Two of these measures, RINe (RIN equivalent) and DV200 (percentage of RNA fragments larger than 200 nucleotides in size), both measured on the Agilent 4200 TapeStation, have become more widely used. The TapeStation offers increased throughput by analyzing 96 samples in a single run (compared to 12 with the Bioanalyzer). Moreover, the manufacturer, Agilent, reports a high correlation between RINe and RIN (r-^2^ = 0.936; median error < ± 0.4 RIN units). In this study, we compared RINe and DV200 to the gold standard RIN using the same set of human brain samples to assess how well the different measurements aligned.

## Materials and Methods

### Samples Preparation

Frozen human tissue samples used in this study were collected as part of the Arizona Study of Aging and Neurodegenerative Disorders (AZSAND) and the Brain and Body Donation Program (BBDP; www.brainandbodydonationprogram.org), a program dedicated to the longitudinal clinicopathological study of neurodegenerative diseases and normal aging (Beach et al., 2015; Beach et al., 2008). All volunteer subjects had signed an Institutional Review Board-approved informed consent for autopsy and post-mortem tissue donation. The median post-mortem interval (PMI) was 3.2 hours.

A total of 183 frozen cerebellum tissue samples from different donors were collected. Qiagen RNeasy Plus mini kits (Qiagen, cat #74134) were used to extract RNA from 25 mg of frozen cerebellum following a previously published protocol (Birdsill, Walker, Lue, Sue, & Beach, 2011; Walker et al., 2016). The tissue was lysed with mild sonication in an RNA-lysis buffer with 0.1M B-Mercapoethanol and processed according to the manufacturer’s instructions. The concentration and purity of each RNA sample were assessed using a Nanodrop Lite spectrometer (Thermo Fisher, cat #ND-LITE). All RNA samples had 260/280 absorbance ratio of at least 2.0, with concentrations ranging from 37.3 ng/uL to 476.3 ng/uL with an average concentration of 228 ng/uL. The RNA was stored at -80C until the time of analysis.

### Instrument Analysis

A volume of 1uL of undiluted RNA from each sample was run in parallel on the Agilent 4200 Tapestation and 2100 Bioanalyzer using the RNA screen tape (Agilent cat # 5067-5576) and RNA 6000 Nano kit (Agilent cat # 5067-1511) respectively. Both instruments require a minimum sample volume of 1ul and the quantitative range for each is 25-500 ng/uL. RIN was recorded from the Bioanalyzer and RINe and DV200 were recorded from the Tapestation 4200. RIN and RINe are assigned to the sample automatically. DV200 is the percent of sample that is above 200 nucleotides and is determined post assay by entering 200 as the lower limit in the regional settings of the Tapestation 4200 software. Details on calculating the DV200 can be found in Illumina’s technical note on Evaluating RNA Quality from Formalin-Fixed Paraffin-Embedded (FFPE) Samples. Both RIN and RINe are expressed as a number between 1 and 10, while DV200 is expressed as a percentage. A cut-off value of RIN 6.5 and DV200 of 70% was used to determine how many samples would be suitable for most downstream applications.

Statistical analyses, including linear regression and paired t-test, were performed using GraphPad Prism v.5 (GraphPad Software, La Jolla, CA) to assess the relationships between RIN, RINe, and DV200 and compare their values. The statistical significance level used for each was p<0.05.

## Results

For all the 183 samples, the RIN range was between 3 to 10 with an average of 8.8 ± 1.06 and a median of 9.1 whereas the RINe range was 2.6 to 7.5 with an average of 5.6 ± 1.07 and a median of 5.8. DV200 ranged from 80.72% - 95.06% with an average of 91.08% ± 2.60% and a median of 91.76% (Figure 1). A two-tailed paired t-test showed that the RIN and RINe were significantly different (p<0.0001) with an average difference of 3.2 RIN units. Linear regression showed that although the RIN and RINe did significantly correlate (p<0.0001), the correlation was weaker than previously reported by Agilent (r^2^ = 0.393 vs 0.936) in their technical note titled “Comparison of RIN and RINe Algorithms for the Agilent 2100 Bioanalyzer and the Agilent 2200 TapeStation systems.” Linear regression comparing RIN to DV200 and RINe to DV200 was also statistically significant (p<0.0001) but with weak correlations of r^2^=0.187 and r^2^=0.346 respectively (Figure 2). Applying a commonly used quality threshold of RIN 6.5, 175/183 (95.6%) samples would be considered fit for downstream applications, whereas only 43/183 (23.5%) would be fit using the RINe and these proportions are statistically significantly different (p <0.0001). Applying a quality cut-off value of 70% DV200, established by Illumina for sequencing, 183/183 (100%) samples would meet this standard (Figure 3).

**Figure 1:**
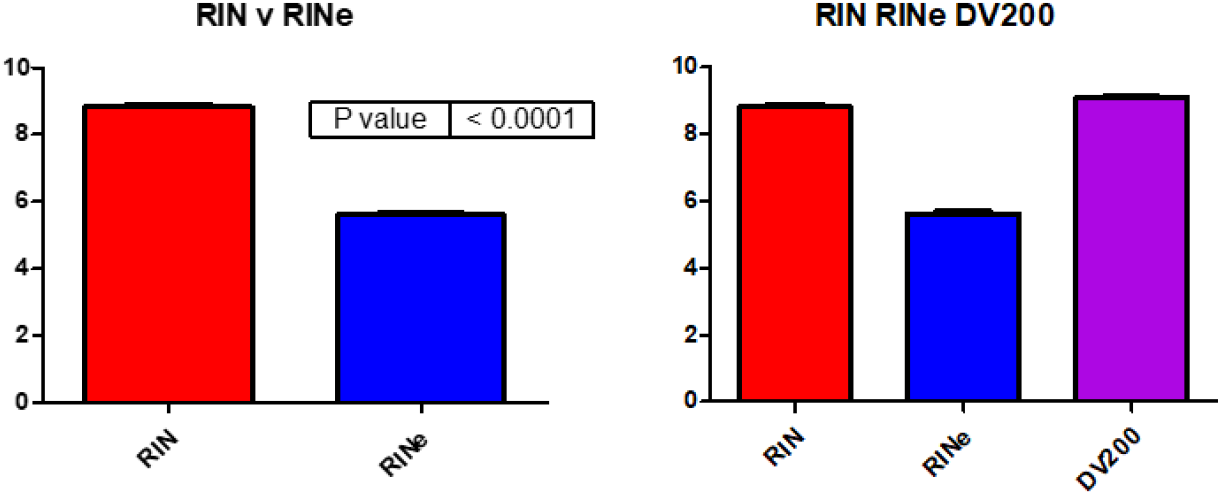
Comparison of RIN, RINe and DV200. For the same 183 human samples, the mean RIN value (Bioanalyzer) was 8.8 ± 1.06 SD with a range of 3-10, and the mean RINe value (TapeStation) was 5.6 ± 1.07 SD with a range of 2.6-7.5. The mean DV200 was 91.08% ± 2.60% SD with a range of 80.7-95.1. DV200 which is expressed as a percentage was modified (100% = 10) for comparison to RIN and RINe.

**Figure 2:**
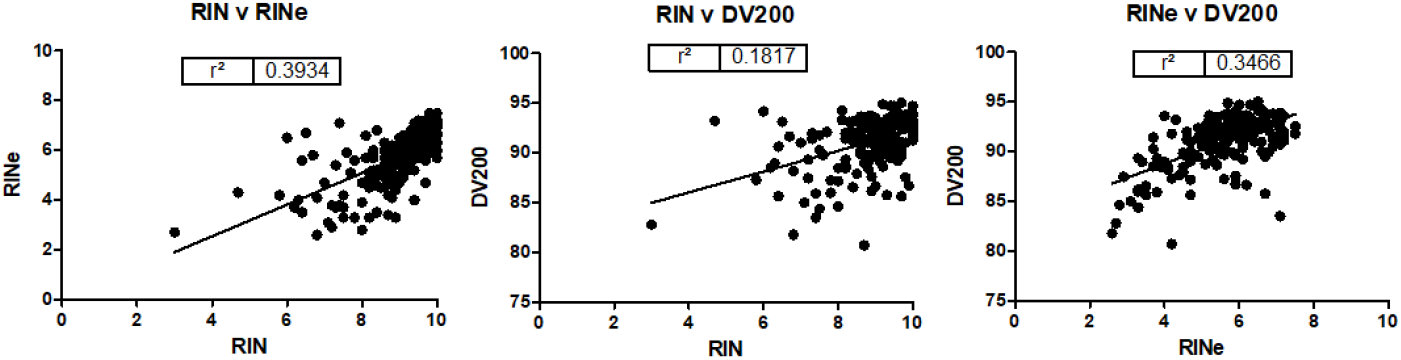
Linear regression comparing the relationship between RIN, RINe, and DV200. All three measurement types-RIN, RINe and DV200 were weakly correlated. RIN vs RINe (r^2^ = 0.3934), RIN vs DV200 (r^2^ = 0.1817) and RINe vs DV200 (r^2^ = 0.3466)

**Figure 3:**
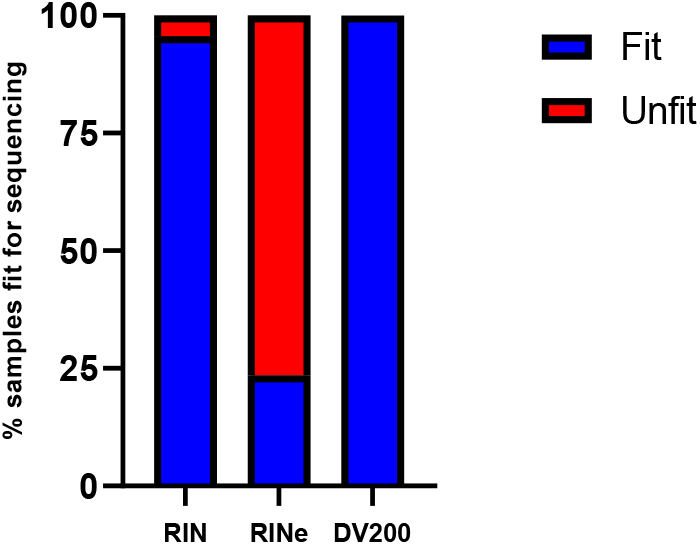
Percent of samples fit for downstream applications per measurement type. Using RIN > 6.5 and DV200 > 70% threshold, the percent of samples that would pass would be 95.6% (175/183) for Bioanalyzer RIN, 23.5% (43/183) for Tapestation RINe and 100% (183/183) for DV200

## Discussion and Conclusion

Concentration, purity, and integrity are all elements to be taken into consideration when evaluating the quality of RNA as they can significantly impact the success or failure of an experiment. It is important to have reliable measurements of RNA quality while also understanding the strengths and limitations of each metric. Absorbance values derived from spectrophotometer (Nanodrop) and fluorometer (Qubit) analysis are common and relatively affordable options for quickly analyzing RNA quality.

The Nanodrop measures the absorbances of the nucleic acids (260 nm) relative to contaminants such as protein (280 nm) or common impurities from RNA purification such as phenol (230 nm). It provides a good estimate of RNA’s concentration and purity. However, the nanodrop does not discriminate between RNA and genomic DNA (gDNA) which also absorbs at 260nm.

The Qubit fluorometer assesses RNA quality with assay Qubit RNA IQ which uses two dyes. One dye binds to large intact RNA and the other binds to small, degraded RNA fragments. The Qubit provides a number between 1-10 as an assessment of quality. Similar to the Nanodrop, the fluorescent dye used by the Qubit is not specific to RNA and can also bind with gDNA present in the sample which may contribute to an overestimate of the RNA concentration. Therefore, while both the Nanodrop and Qubit are valuable tools for assessing RNA quality, it is important to be aware of their limitations, especially regarding the potential interference from gDNA when interpreting results.

Samples can be analyzed via gel electrophoresis allowing the 28S:18S ribosomal RNA ratio to be determined with visualization of both gDNA contamination and degraded RNA. Typically, a 28S:18S ratio of 2:1 is considered indicative of good quality (Imbeaud et al., 2005). However, these ratios must be subjectively interpreted, making them dependent on inter-rater reliability. The TapeStation and Bioanalyzer are based on the gel electrophoresis method and aim to standardize this approach. In both the Tapestation and Bioanalyzer, as RNA degrades, the 28S and 18S peaks become less pronounced and smaller/degraded peaks become more prevalent leading to a reduction in the quality score.

Although the instruments appear similar in their analysis, they are ultimately based on different algorithms. The Tapestation measures the relative ratio of the degraded products in the fast zone to the 18S peak signal where the fast zone is defined as the region between small RNAs and the 18S. As total RNA degrades, the 18S and 28S rRNA peaks slowly disappear and degradation products emerge in the fast zone. On the other hand, the Bioanalyzer assesses the entire electrophoretic trace including the 28S and 18S peak ratios, their separation, and the presence or absence of degraded products.

Our results suggest that there may be larger discrepancies between RIN and RINe than previously reported, emphasizing that these measurements should not be used interchangeably. Instead, separate quality thresholds should be established for each metric. This is an important observation because currently the broader scientific community may be working under the assumption that these methods are virtually equivalent. A general premise in the field is that RIN values lower than 6.5 are not considered adequate for downstream applications. If the same standards were applied to RINe, many RINe samples will fail quality control (QC) that would pass with RIN. This was seen in our study where only 23.5% of samples would pass QC using RINe but 95.6% of samples would pass QC using RIN.

Several explanations may be put forward to explain discrepancies between our study and previously published results. The manufacture’s comparison of RIN and RINe included commercially prepared cell lines as well as RNA from humans, rats, and mice. They also degraded total RNA by heating for various durations (0 to 120 minutes) to compare samples with lower RNA integrity. RIN values ranged from 1.2 to 10. These methods are further described in Agilent’s Technical Overview “Comparisons of RIN and RINe Algorithms for the Agilent 2100 Bioanalyzer and the Agilent 2200 Tapestation System”. In our comparison of RIN and RINe, we only analyzed RNA from postmortem human brain samples and did not deliberately degrade the RNA to expand the range of RNA integrity; therefore, we had few samples with poor RNA integrity to compare. In this study, the highest values obtained for RIN extended up to the maximum of 10, while the maximum RINe obtained was 7.5. We hypothesize that the type of degradation that occurs due to normal postmortem processing delays is different than deliberate heat denaturation after RNA Isolation that Agilent performed, and this could explain some of the differences in results. It is also possible that the Tapestation’s algorithm is more sensitive to the type of degradation that occurs during normal postmortem processing while their reported high values of RINe 9 and 10 were achieved with commercially prepared cell lines.

Considering that RINe continues to gain popularity as a quality assessment, more research still needs to be done to better assess what constitutes an appropriate quality threshold of this measure. One recent study indicated that DV200 is a better predictor than RINe for Next Generation Sequeincing (NGS) success when both measurements were correlated with NGS library yields (Matsubara et al., 2020). This was especially true with low-quality RNA samples derived from FFPE samples. In addition to being a better predictor of success, approximately 37.5% of samples with low RINe < 5 had a DV200 of above 70%, indicating that RINe may exclude many adequate samples for a sequencing study. To date, no similar end-use fitness studies have been done using Bioanalyzer-determined RIN. In this study, we only report a weak correlation between DV200 and RIN or RINe. However, when these metrics are independently used to evaluate the likelihood of success in downstream applications, RIN and DV200 agreed on 95% of cases while RINe and DV200 agreed only on 23% of cases.

## Data Availability

All data produced in the present study are available upon reasonable request to the authors

## Acknowledgments

The Arizona Study of Aging and Neurodegenerative Disorders and Brain and Body Donation Program has been supported by the National Institute of Neurological Disorders and Stroke (U24 NS072026 National Brain and Tissue Resource for Parkinson’s Disease and Related Disorders), the National Institute on Aging (P30 AG19610 and P30AG072980, Arizona Alzheimer’s Disease Center), the Arizona Department of Health Services (contract 211002, Arizona Alzheimer’s Research Center), the Arizona Biomedical Research Commission (contracts 4001, 0011, 05-901 and 1001 to the Arizona Parkinson’s Disease Consortium), and the Michael J. Fox Foundation for Parkinson’s Research.

The authors thank the autopsy personnel who helped contribute clinical data and postmortem brains from study subjects, and the donors who were recruited for this study as well as their families.

